# Acute myeloid leukemia with mixed phenotype is characterized by *RUNX1* mutations, stemness features and limited lineage plasticity

**DOI:** 10.1101/2023.11.01.23297696

**Authors:** Pallavi Galera, Deepika Dilip, Andriy Derkach, Alexander Chan, Yanming Zhang, Sonali Persuad, Tanmay Mishera, Ying Liu, Christopher Famulare, Qi Gao, Douglas A. Mata, Maria Arcila, Mark B. Geyer, Eytan Stein, Ahmet Dogan, Ross L. Levine, Mikhail Roshal, Jacob Glass, Wenbin Xiao

## Abstract

Mixed phenotype (MP) in acute leukemias poses unique classification and management dilemmas and can be seen in entities other than de novo mixed phenotype acute leukemia (MPAL). Although WHO classification empirically recommends excluding AML with myelodysplasia related changes (AML-MRC) and therapy related AML (t-AML) with mixed phenotype (referred to as “AML-MP”) from MPAL, there is lack of studies investigating the clinical, genetic, and biologic features of AML-MP. We report the first cohort of AML-MP integrating their clinical, immunophenotypic, genomic and transcriptomic features with comparison to MPAL and AML without MP. Patients with AML-MP share similar clinical and genetic features to its AML counterpart but differs from MPAL. AML-MP harbors more frequent *RUNX1* mutations than AML without MP and MPAL. *RUNX1* mutations or complex karyotypes did not impact the survival of MPAL patients. Unsupervised hierarchal clustering based on immunophenotype identified biologically distinct clusters with phenotype/genotype correlation and outcome differences. Furthermore, transcriptomic analysis showed an enrichment for stemness signature in AML-MP and AML without MP as compared to MPAL. Lastly, MPAL but not AML-MP often switched to lymphoid only immunophenotype after treatment. Expression of transcription factors critical for lymphoid differentiation were upregulated only in MPAL, but not in AML-MP. Our study for the first time demonstrates that AML- MP clinically and biologically resembles its AML counterpart without MP and differs from MPAL, supporting the recommendation to exclude these patients from the diagnosis of MPAL. Future studies are needed to elucidate the molecular mechanism of mixed phenotype in AML.

**Key points:** AML-MP clinically and biologically differs from MPAL but resembles AML.

AML-MP shows *RUNX1* mutations, stemness and limited lineage plasticity.

## INTRODUCTION

Mixed phenotype, defined by the presence of two or more lineage distinct blast populations or a single blast population with lineage ambiguity/promiscuity, is characteristic for de novo mixed phenotype acute leukemia (MPAL) but can also be rarely seen in other types of acute leukemia. Specifically, AML with myelodysplasia-related changes (AML-MRC) and therapy-related AML (t-AML) can manifest as mixed phenotype (referred to as AML-MP), which poses unique classification and management dilemmas. Although the WHO classification of hematolymphoid tumors (2017) empirically recommends excluding AML-MP from the diagnosis of MPAL, ^1,2^ there is a complete lack of studies investigating the clinical, genetic and biologic features of AML-MP except a few single case reports. ^3,4^ Studies have shown that regimens directed towards acute lymphoblastic leukemia (ALL) may result in better outcome in patients with MPAL;^5–10^ whereas, liposomal formula of cytarabine and daunorubicin has become the standard of care for patients with AML-MRC and t-AML.^11^ Therefore, a better delineation between AML-MP and MPAL is urgently needed for clinical decision making.

Genetically, MPAL often harbor fusions/alterations involving *BCR::ABL1*, *KMT2A*, *ZNF384*, *BCL11B* and *PICAML::MLLT10,*^12^ and mutations in *PHF6*, *DNMT3A*, *NOTCH1* and *WT1*. ^13–17^ In contrast, AML-MRC is enriched for myelodysplasia (MDS)-defining cytogenetic abnormalities (MDS-CG) and so-called “MR” gene mutations (i.e., *SRSF2, SF3B1, U2AF1, ZRSR2, ASXL1, EZH2, BCOR*, and *STAG2* with or without *RUNX1*).^18,19^ *TP53* mutations and monosomal/complex karyotypes are frequent in t-AML.^20^ The genetic landscape of AML-MP remains elusive. Furthermore, the newly proposed WHO2022 and ICC2022 classifications^12,21^ of hematolymphoid tumors both include MR gene mutations as one of the diagnostic criteria for AML-MR. ICC2022 (but not WHO2022) further includes *RUNX1* mutations to the list of MR genes while removes t-AML as an entity. These drastic changes create uncertainties in the diagnostic algorithm of MPAL. Although WHO2022/ICC2022 both recommend classifying acute leukemia with mixed phenotype and *MR/RUNX1* mutations as AML-MR, there is scarce data to verify it. Relatedly, it is uncertain whether therapy-related history still matters or not^22^.

In this study, we report the first cohort of AML-MP to this date integrating the clinical, immunophenotypic, genetic, and transcriptomic features, with comparison to MPAL and AML without MP. We demonstrate that 1) The clinical, genetic, and transcriptomic features of AML-MP closely resemble that of AML without MP and differ from that of MPAL; 2) AML-MP has frequent *RUNX1* mutations and is enriched for stemness signatures; 3) *RUNX1* mutations (with or without MR) and complex karyotype do not impact the outcome of MPAL; 4) AML-MP, despite mixed phenotype, has limited lymphoid lineage plasticity compared to MPAL.

## METHODS

### Patient cohorts

The MSKCC pathology database was searched from January 2014 to December 2022 for a new diagnosis of “acute leukemia” or “acute myeloid leukemia” and/or “mixed phenotype”. A second search was performed on flow cytometry reports with “abnormal myeloid blast” and/or “abnormal immature T cells” and/or “abnormal immature B cells”. The definition of mixed phenotype was based on the WHO Classification of Haematolymphoid Tumours 2017.^2^ In this study, flow cytometric evaluation was used, and the lineage assignment was independently confirmed by at least 2 of the participating board-certified hematopathologists (P.G., A.C., M.R. and W.X.). Another cohort of 100 patients newly diagnosed as AML- MRC or t-AML without MP was also included for comparison. The clinical, morphologic, immunophenotypic, cytogenetic and molecular results of all patients at initial diagnosis were manually re- annotated. This study was approved by the institutional review board (IRB) at MSKCC, and informed consents were obtained from all patients.

Detailed methods including chromosome and FISH analysis, flow cytometry (Table S1) and cell sorting, RNA sequencing and statistical analysis are included in supplemental data. For original data, please contact.

## RESULTS

### Clinical characteristics of AML-MP

A total of 137 newly diagnosed patients with ≥20% blasts and mixed phenotype were initially identified and confirmed. Patients with a diagnosis of blast phase of chronic myeloid leukemia (CML) (18, 13%), blast phase of *BCR::ABL1* negative myeloproliferative neoplasms (MPN) (3, 2.2%), B-lymphoblastic leukemia/lymphoma (B-ALL) with isolated myeloperoxidase expression (7, 5.1%), AML with recurrent genetic abnormalities (7, 5.1%), and myeloid/lymphoid neoplasms with eosinophilia and rearrangements (2, 1.5%) were excluded (**Figure 1A**). The remaining 55 (40%) patients of AML-MP (40 AML-MRC and 15 t-AML) and 45 (33%) patients of MPAL were included in this study (**Figure 1A and Table S2**). Therefore, only a third of acute leukemia with mixed phenotype is *bona fide* MPAL. Of note, 26 out of 40 (65%) AML-MRC with MP had no history of MDS or MDS/MPN and the diagnosis was based on the presence of MDS-CG (**Figure 1B**). One patient with neither history nor MDS-CG had a mutational profile (*ASXL1, RUNX1, SRSF2*, three *TET2*, two *STAG2, CEBPA, FLT3*, and *MED12*) typical for AML and therefore was eventually classified as AML-MP after group consensus. The overall frequency of mixed phenotype in AML-MRC and t-AML was 3.3% (55/1679) with 2.8% (40/1429) in AML-MRC and 6% (15/250) in t-AML, respectively.

**FIGURE 1:**
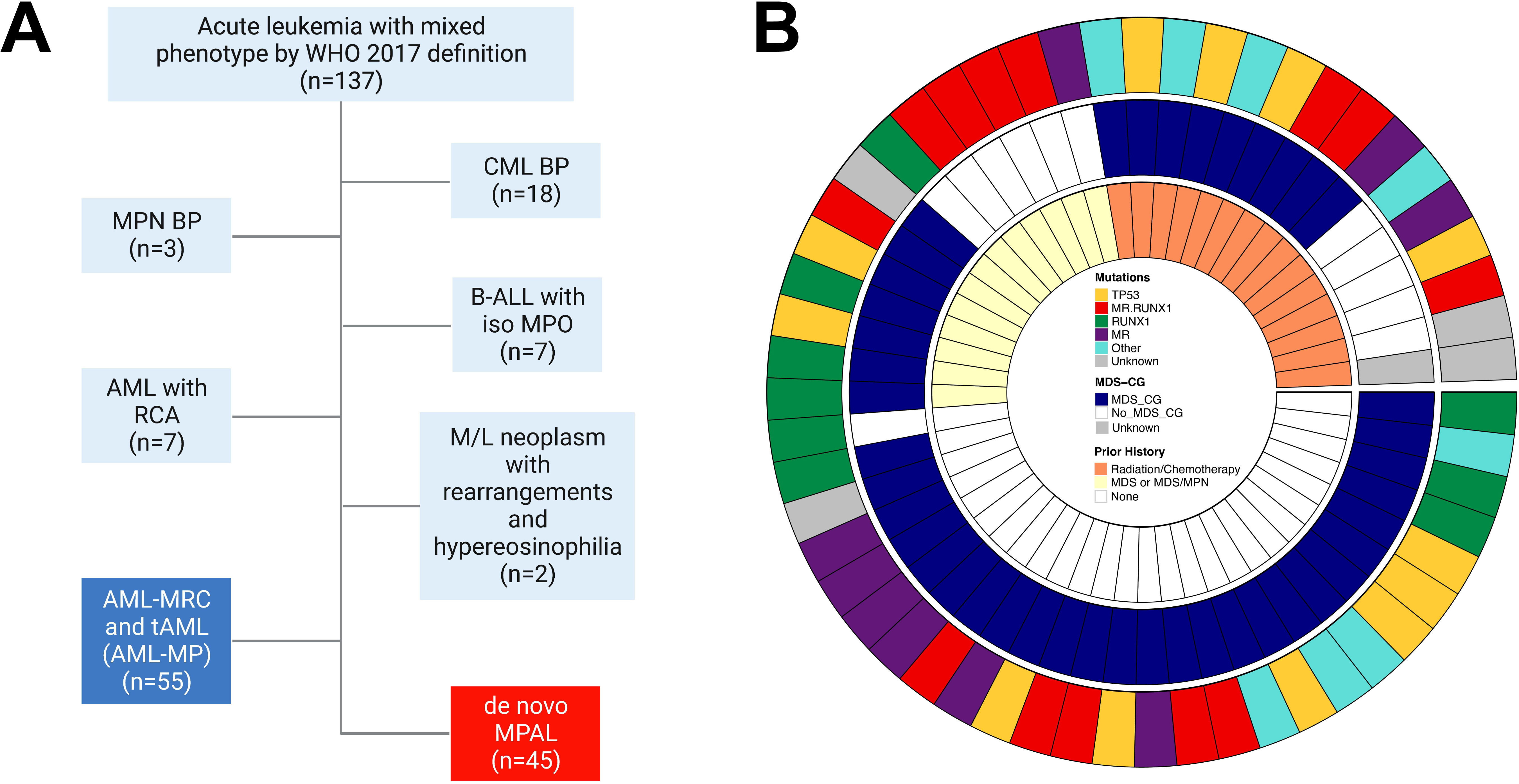
Diagnostic algorithm for AML-MP and MPAL. **A.** Inclusion and exclusion criteria of AML-MP and MPAL cohorts. Only 1/3 of patients presented with acute leukemia with mixed phenotype had bona fide MPAL. Abbreviations: CML BP, chronic myeloid leukemia with blast phase; MPN BP, myeloproliferative neoplasm with blast phase; B-ALL with iso MPO, B-acute lymphoblastic leukemia with isolated myeloperoxidase expression; AML with RCA, acute myeloid leukemia with recurrent cytogenetic abnormalities; M/L, myeloid and lymphoid; AML-MRC, acute myeloid leukemia with myelodysplasia- related changes; t-AML, therapy-related AML; MPAL, mixed phenotype acute leukemia. **B.** Pie chart of the AML-MP cohort detailing the inclusion criteria namely prior history of cytotoxic chemotherapy and/or radiotherapy administered for a non-myeloid neoplastic or non-neoplastic disorder for t-AML (15 cases) and prior history of MDS and/or MDS/MPN and/or the presence of MDS defining cytogenetic abnormalities for AML-MRC (39 cases; 6 cases with prior history of MDS and/or MDS/MPN only, 26 cases with MDS defining cytogenetic abnormalities only, and 7 cases with both). 1 case lacking both history of MDS and/or MDS/MPN and MDS defining cytogenetic abnormalities had mutational profile typical for AML and was classified as AML-MRC. MR.RUNX1 represents patients with *MR* and *RUNX1* comutations. MR represents patients with *MR* mutations (not including *RUNX1*). RUNX1 represents patients with *RUNX1* mutations while no other *MR* mutations. MR mutations include mutations in any of these genes: *ASXL1, BCOR, EZH2, STAG2, SF3B1, SRSF2, U2AF1,* and *ZRSR2.* MDS-CG, MDS related cytogenetic abnormalities which included complex karyotype (≥ 3 abnormalities), 5q deletion or loss of 5q due to unbalanced translocation, monosomy 7, 7q deletion, or loss of 7q due to unbalanced translocation, 11q deletion, 12p deletion or loss of 12p due to unbalanced translocation, monosomy 13 or 13q deletion, 17p deletion or loss of 17p due to unbalanced translocation, isochromosome 17q and idic(X)(q13)^12^. Abbreviations: MDS, myelodysplastic syndrome/neoplasm; MDS/MPN, myelodysplastic and myeloproliferative overlap syndrome.

We compared the clinical features of AML-MP to that of AML without MP (63 AML-MRC and 37 t-AML) and MPAL (**Table 1**). The median age at diagnosis of AML-MP was 66.5 years, significantly higher than that of MPAL (42 years, p<0.0001) but similar to that of AML without MP (70 years). All 3 cohorts had male preponderance. Both AML cohorts (with or without MP) had fewer peripheral/bone marrow blasts but more severe leukopenia and thrombocytopenia than MPAL. The AML-MP cohort included 30 (54.5%) B/Myeloid (B/M), 20 (36.4%) T/M and 5 (9.1%) B/T/M phenotype, similar to MPAL [26 (57.8%), 16 (35.6%), and 2 (4.4%), respectively]. One patient with MPAL had a rare T/B phenotype. Fourteen (25.5%) cases of AML-MP revealed extramedullary involvement (7 involving skin and 7 lymph node), higher than that seen in AML without MP cases (8%, 5 skin and 3 lymph node involvement, p=0.007). While the MPAL cohort demonstrated similarly high numbers of cases with extramedullary involvement (12/45, 26.7%) the distribution was quite different from AML-MP with nearly all MPAL patients revealing nodal and only 1 patient showing skin involvement (p=0.03, **Table S3**).

**Table 1.**
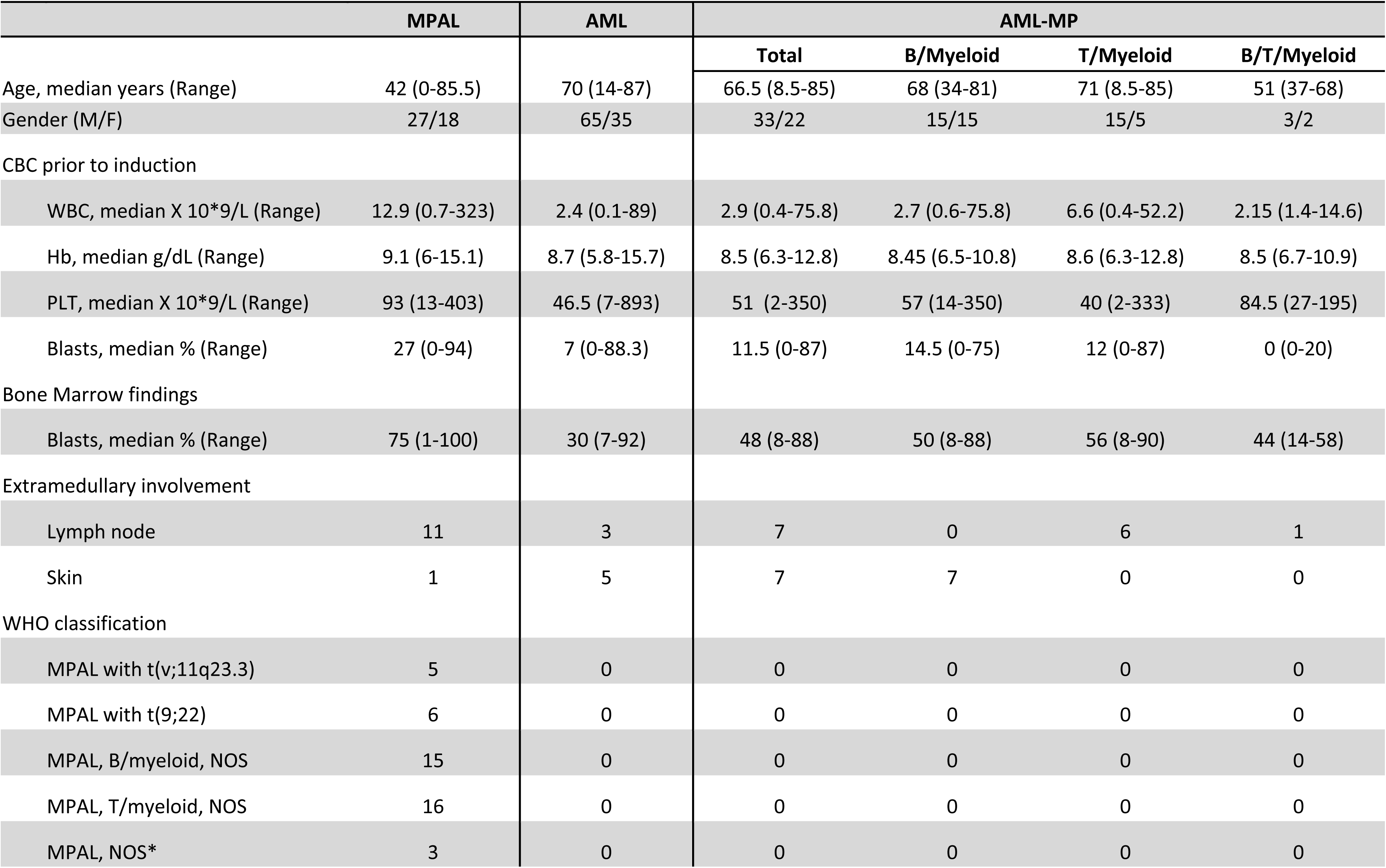
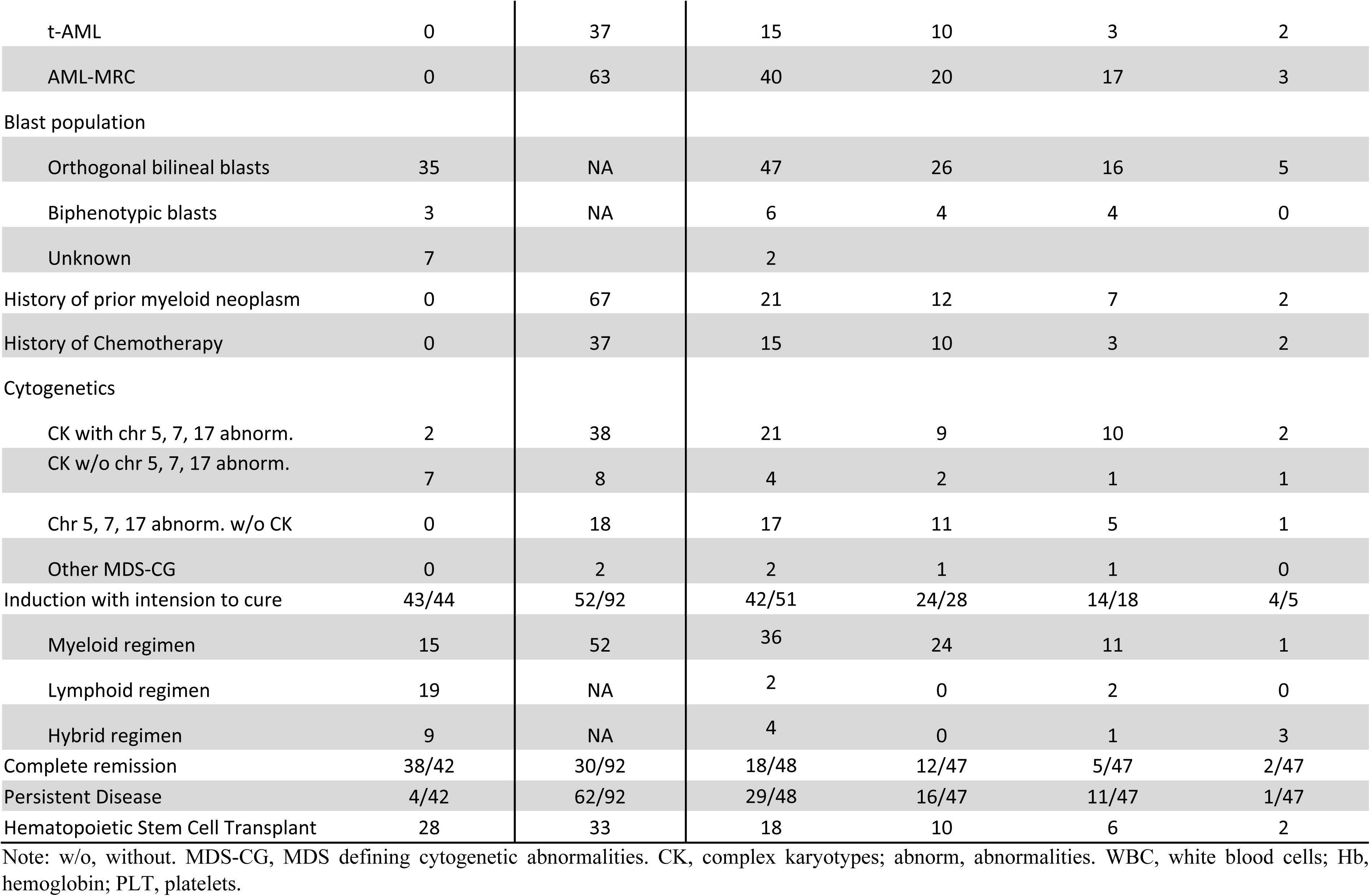
Clinicopathological Characteristics of patients with AML-MP and MPAL.

### Genomic landscape of AML-MP

Mutational profiles were revealed by targeted NGS sequencing studies in 52 of 55 AML-MP, 37 of 45 MPAL and 99 of 100 AML without mixed phenotype (**Figure 2**). Mutations in *TET2, FLT3*, *KRAS*, *PTPN11*, *IDH1*, *IDH2*, and *CEBPA* showed no statistical difference among the 3 cohorts. Notably, mutations in *TP53* were absent from MPAL. Although MR gene coverage was limited in most patients, *SRSF2*, *U2AF1*, *BCOR*, *EZH2*, *STAG2* and *SETBP1,* while variably common in both AML cohorts, were notably absent from MPAL. Mutations in other MR genes such as *ASXL1* and *SF3B1* were also rare in MPAL. Conversely, MPAL was enriched for mutations in *PHF6* and fusions in *MLL*, *BCR::ABL1* and *PICALM::MLLT10*. *RUNX1* mutations, although common in all 3 cohorts, were significantly more frequent in AML-MP [21/52 (40.4%) vs 23/99 (23%) in AML without MP, p=0.02 and 7/35 (20%) in MPAL, p=0.06]. *NRAS* mutations were also more frequently present in AML-MP (15.7% vs 4% AML without MP, p=0.02). In contrast, mutations in *SRSF2*, *U2AF1* and *SETBP1* were more common in AML without MP than AML-MP (**Figure 2B**).

**FIGURE 2:**
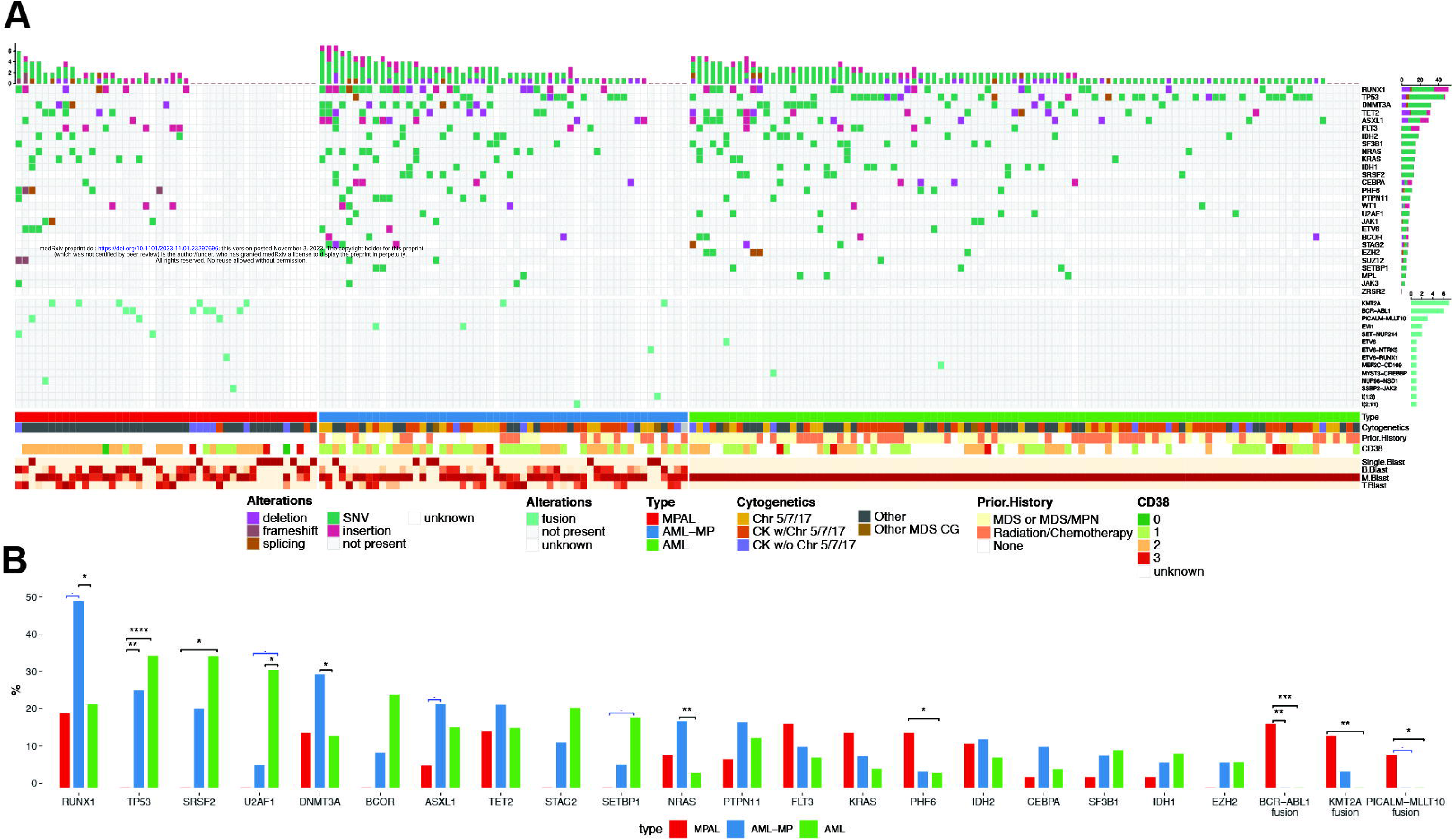
Landscape of gene mutations and cytogenetic abnormalities in AML-MP and MPAL. **A.** The oncoplot tabulates mutations, fusions, cytogenetics, prior history of cytotoxic chemotherapy and/or radiotherapy, prior history of MDS and/or MDS/MPN for all the 3 cohorts (AML-MP, MPAL and AML without MP) along with the blast lineage and CD38 expression intensity annotated on the bottom bar. The genes not included in the NGS panel during sequencing were marked as blank. B blasts, B-lineage blasts; M blasts, myeloid lineage blasts; T-blasts, T-lineage blasts. SNV, single nucleotide variant. Chr5/7/17, MDS-cytogenetic abnormalities involving chromosomes 5, 7 or 17 but not qualifying for complex karyotype; CK w/ Chr5/7/17, complex karyotype with MDS defining cytogenetic abnormalities involving chromosomes 5, 7 or 17; CK w/o Chr5/7/17, complex karyotype but not involving chromosomes 5, 7 or 17. **B**. Bar graphs showing pairwise comparisons of mutation and fusion frequencies within the 3 cohorts, containing statistical significance bars in black and trend towards significance in blue. *p<0.05, **p<0.01, ***p<0.001.

MDS-CG was frequent in AML-MP (80%) and AML without MP (66%) (**Table 1**). In contrast, only 9/45 (20%) patient with MPAL had complex karyotypes (CK, **Table S4)**: 1 had a T/B (no myeloid) phenotype, 1 had a *PICALM-MLLT10* translocation with isochromosome 17q(10), 5 had other balanced translocations and CK not involving chr. 5, 7 or 17, and finally 2 had a hyper-diploid clone (1 with a sub-clonal del5q). Thus, despite of atypical complex karyotypes, these 9 patients were classified as MPAL.

### Unsupervised hierarchal clustering based on immunophenotyping

To explore the immunophenotype in an unbiased manner, raw clinical flow cytometry data in the form of FCS files were computationally analyzed in an unsupervised fashion (see Supplemental data). The viable, dim CD45 expressing cells were automatedly extracted to loosely capture leukemic blasts. 36 distinct populations derived from M1 and M2 tubes were identified in all the samples from 3 cohorts using a self- organizing map (SOM) (**Figure S1-S3**). Hierarchical clustering using these populations revealed 8 distinct clusters of patients (**Figure 3**). Most clusters exhibited a relatively uniform lineage and composition of either AML without MP, AML-MP or MPAL, but some were more variable suggestive of either suboptimal separation related to limited protein markers available or a potential biological similarity among subsets of these cohorts.

**FIGURE 3:**
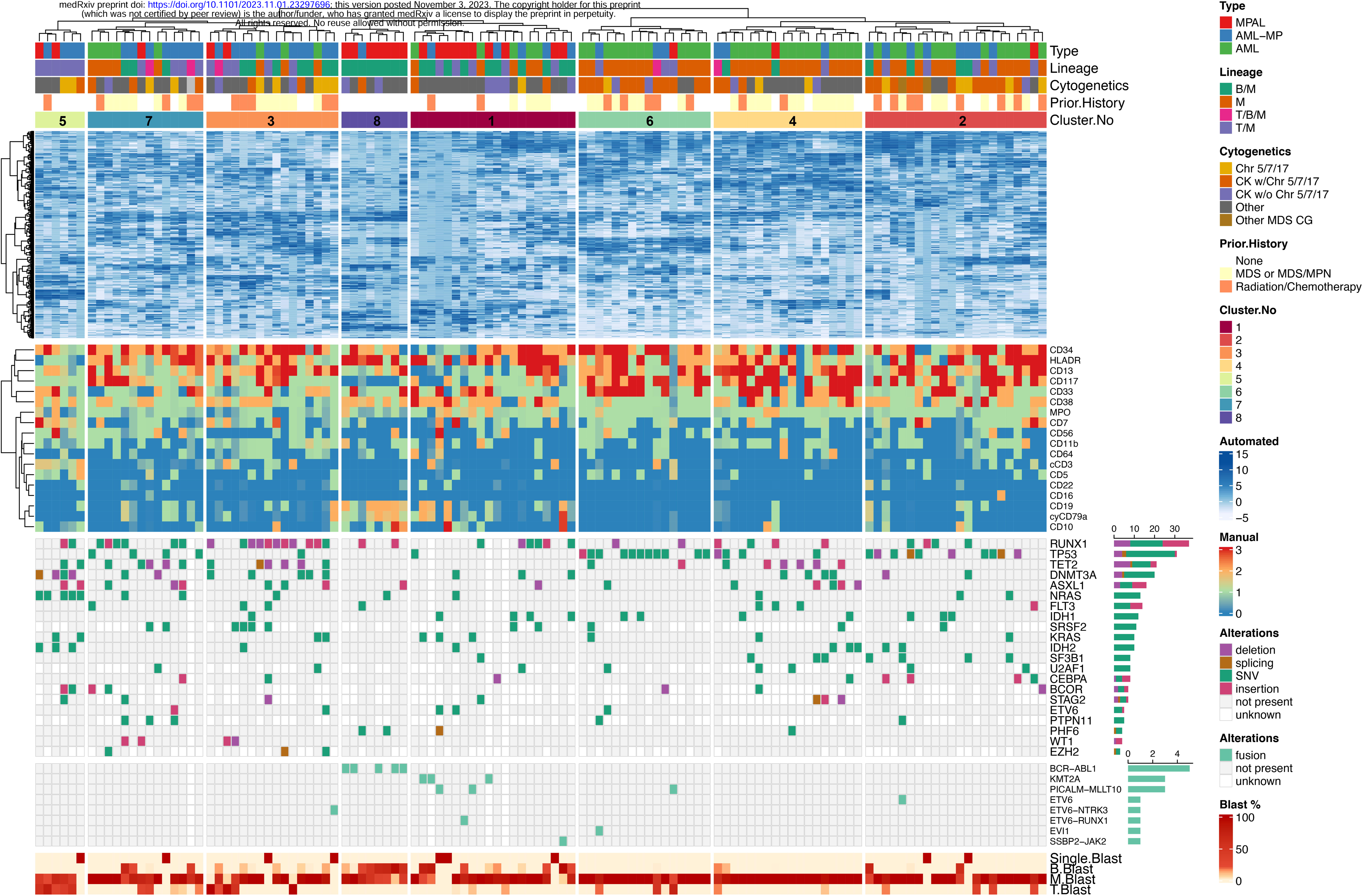
Unsupervised hierarchal clustering based on automated immunophenotypic data of AML-MP and MPAL. Hierarchical clustering (Euclidean distance, Ward’s method) was used to create eight distinct groups of patients based on automated analysis on raw clinical flow cytometry data in the form of FCS files of M1 and M2 tubes from the 3 cohorts (see Supplemental Table S1). Disease type, blast lineage, cytogenetics, and prior history were listed on top of the plot. The cell populations identified by automated analysis were shown at the upper portion of the plot. The automated analysis was contrast with manually annotated immunophenotype as indicated by CD markers (middle portion of the plot). Additionally, mutations and fusions were listed at the lower portion of the plot. Blast ratio was listed at the bottom of the plot.

To better understand the biology driving the clustering, disease type, cytogenetic changes, molecular alterations and manually annotated immunophenotype were evaluated for enrichment within each cluster. A B/Myeloid mixed phenotype were primarily observed in Clusters 1 (10/20, 50%), 3 (7/16, 43.8%), and 8 (8/8, 100%). Cluster 3 consisted of AML-MP cases with B/Myeloid mixed phenotype and an enrichment in *RUNX1* mutations (11/16, 69%) with high levels of CD34 and CD13 expression. Clusters 8 represented MPAL with B/Myeloid phenotype with significantly lower levels of CD117 expression (p=0.005). Cluster 8 had more frequent *BCR::ABL1* fusions (5/8, 62.5%), and cluster 1 had various other fusions. Cluster 6 was composed almost entirely of patients with AML with or without MP, mostly harboring *TP53* mutations and a monosomal and/or complex karyotype (11/16, 68.8%). Cluster 6 also had high levels of CD34 and CD117 expression but very low levels of CD38 expression, an immunophenotype enriched for hematopoietic/leukemic stem cells ^23,24^. Despite the lack of T-cell markers in our FCS files, this approach identified cluster 5 with a T/M phenotype (6/6, 100%) and enrichment for mutations in *DNMT3A*, *NRAS* and *IDH2*,^14^ composed of predominantly AML-MP with T/M phenotype and a few MPAL with T/M phenotype, suggesting some higher dimensional immunophenotypic features may be only appreciated by higher dimensional computational analysis. Cluster 4 consisted predominantly of patients with a prior history of MDS/MPN (13/18, 72.2%) and mostly without MP (17/18, 94.4%). The majority of cases in Cluster 2 were AML without MP (13/22, 59%). Altogether, these data demonstrate that immunophenotype based hierarchical clustering may reveal disease biology linking phenotype and genotype.

### AML-MP has similar outcomes to AML without MP but inferior to MPAL

The overall survival (OS) was inferior in patients with AML-MP in comparison to MPAL (median: 10.3 vs 42.8 months, p<0.0001; **Figure 4A**) but comparable to those AML without MP (median: 7.1 months, p=0.3; **Figure 4A**). AML-MP patients were also less likely to achieve complete remission after induction in comparison to MPAL (p<0.001, Table 1). In a multivariable analysis, after adjusting for the set of major confounders such as age, treatment with intension to cure, cytogenetic risk stratification category^25^ and allotransplant, MPAL was still significantly associated with better OS (**Figure 4B**). We also studied the association between unsupervised hierarchical clustering and the outcome. Both clusters 1 and 8 were enriched for MPAL with B/M phenotype, however, cluster 8 had higher ratio of *BCR::ABL1* translocations and showed superior OS (**Figure 4C**). Conversely, cluster 6, characterized by *TP53* mutations had inferior OS. Surprisingly, cluster 5, enriched for T/M phenotype and composed of a mixture of AML-MP and MPAL also had adverse OS. Other clusters showed intermediate OS. Association between 8 clusters and OS remained significant after adjusting for the same set of confounders (p=0.007).

**FIGURE 4:**
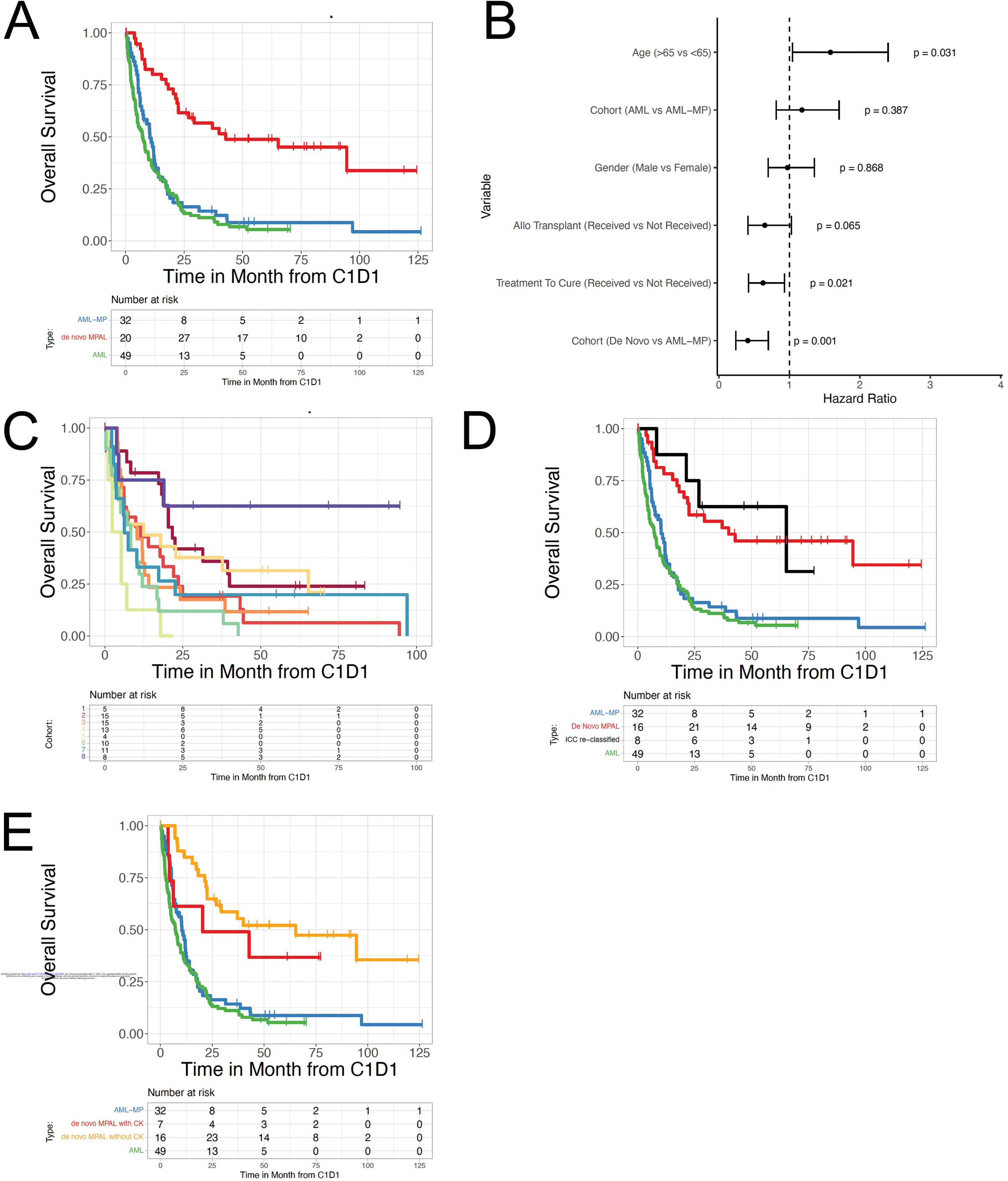
Outcome of AML-MP and MPAL. **A.** Kaplan-Meier curves of overall survival stratified by disease types. Red line, MPAL; green line, AML without mixed phenotype; blue line, AML-MP. **B**. A forest plot indicating hazards ratios per clinical predictors including age, treatment with intension to cure (TreatmentToCures), cytogenetic risk stratification category and allotransplant (INTOallo received). **C.** Kaplan-Meier curves of overall survival stratified by immunophenotypic clusters generated unsupervised hierarchal clustering (Figure 3). **D.** Kaplan-Meier curves of overall survival after re-classifying MPAL cases according to the presence of *RUNX1 (*with or without *MR)* gene mutations. Black line, MPAL patients harboring *RUNX1* (with or without *MR*) mutations; red line, MPAL patients with no *RUNX1* mutations. **E.** Kaplan-Meier curves of overall survival after stratifying MPAL cases according to the presence or absence of complex karyotype. Yellow line, MPAL patients with complex karyotypes; red line, MPAL patients without complex karyotypes.

### *MR/RUNX1* gene mutations or complex karyotypes do not confer inferior outcome to MPAL

As both WHO2022 and ICC2022 classifications^12,21^ expand the AML-MR category by including patients harboring *MR/RUNX1* gene mutations regardless of history and/or cytogenetics, we investigated if the newly included *MR/RUNX1* gene mutations in otherwise MPAL would confer an inferior outcome and thus would suggest reclassifying it as AML-MP. Eight patients of MPAL harbored *MR/RUNX1* gene mutations: 1 *RUNX1/ASXL1*, 1 *RUNX1/SF3B1*, 1 *ASXL1* and 5 *RUNX1*. Interestingly, these 8 patients had similar OS to the remaining MPAL patients but superior to AML-MP (median survival: 65 vs 10 months, p=0.014; **Figure 4D**). The subgroup of patients (n=3) with WHO2022 *MR* gene mutations was too small for outcome analysis.

To examine if complex karyotypes are associated with inferior outcome in MPAL, overall survival was compared between MPAL patients with or without complex karyotype (**Figure 4E**), which showed no difference. Therefore, complex karyotype may not confer inferior outcome in *bona fide* MPAL patient after excluding AML-MP.

### AML-MP shares upregulated stemness gene sets with AML without MP

Gene-expression analysis was performed on flow sorted myeloid- vs T-lineage leukemic blasts using RNA- seq to compare the genetic programs differentiating MPAL from AML-MP cases. Principal component analysis revealed two distinct lineage-based groups: one primarily consisting of MPAL samples and the other with both AML without MP and AML-MP (**Figure 5A**). Interestingly, myeloid- and T-lineage blasts from the same patient clustered closely. This was further confirmed when unsupervised clustering was performed on RNA-seq data, resulting in two groups akin to the PCA (**Figure S4**). Flow marker annotations indicated higher CD38 and lower CD34 levels in the MPAL cluster compared to the AML-MP/AML group, suggesting a more differentiated immunophenotype of MPAL (**Figure S4**).

**FIGURE 5:**
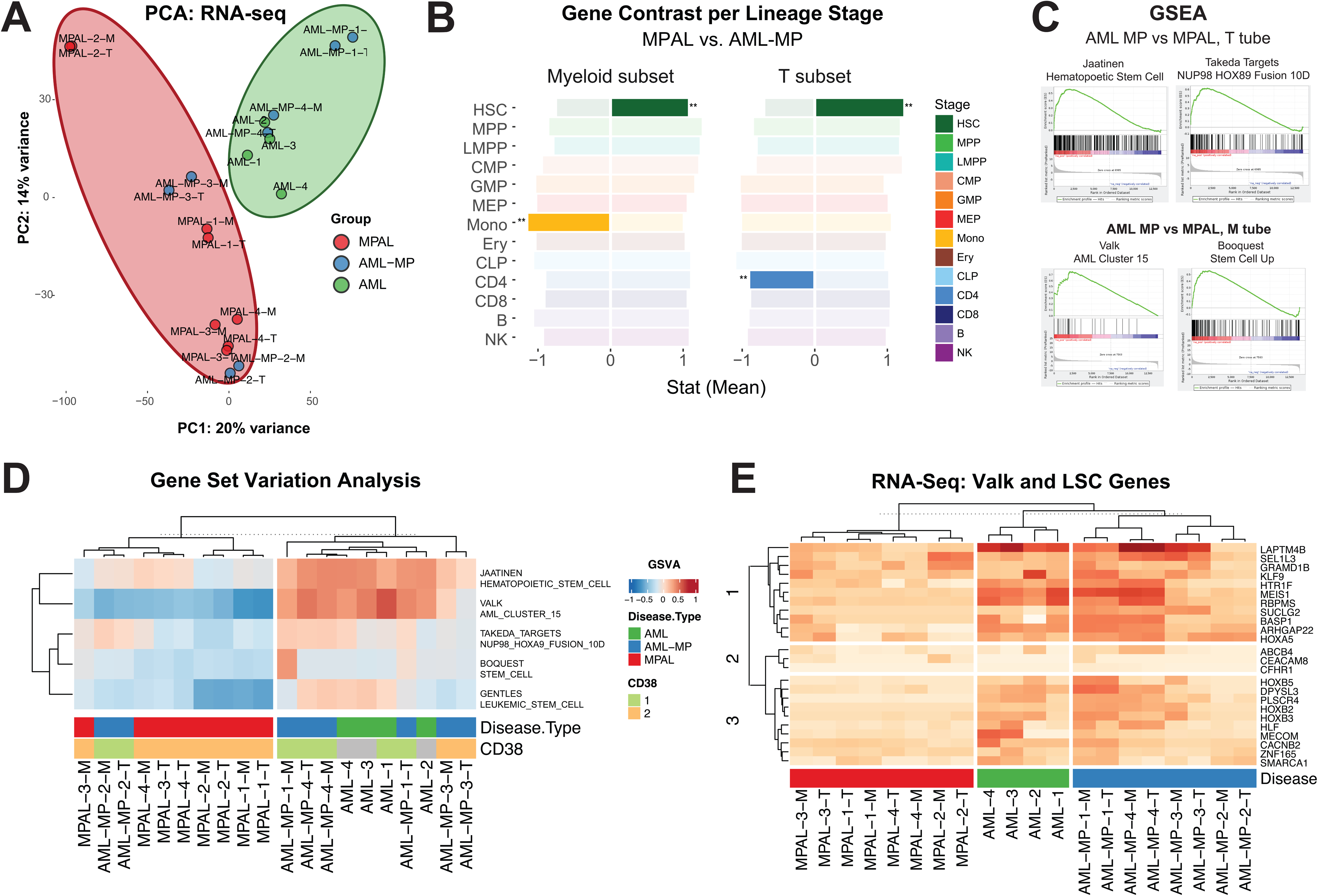
Enriched stemness transcriptomic signatures in AML-MP. **A.** Principal components analysis (PCA) conducted on RNA-seq of flow sorted leukemic blasts, colored by disease type. **B.** Gene set enrichment tests on transcriptome of flow sorted leukemic blasts comparing AML-MP and MPAL cases. The mean statistic for gene contrasts (in both positive and negative directions) is displayed per stage, with statistical significance (p= < 0.05) of the enrichment test indicated. Enrichment was conducted using contrived gene sets per hematopoietic lineage stage, indicating high HSC enrichment in AML-MP samples and high monocytic and CD4 positive T-cell lineage enrichment in myeloid and T- blasts in MPAL samples. HSC, hematopoietic stem cells; MPP, multipotent progenitors; LMPP, lymphoid myeloid potent progenitors; CMP, common myeloid progenitors; GMP, granulocytic and monocytic progenitors; MEP, megakaryocytic and erythroid progenitors; CLP, common lymphoid progenitors; Ery, erythroid; NK, nature killer cells; CD4, CD4 positive T cells; CD8, CD8 positive T cells; B, B cells. **C.** Gene set enrichment analysis (GSEA) comparing AML-MP and MPAL cases, indicating stem cell enrichment in AML-MP. **D.** Gene set variation analysis (GSVA) clustered using hierarchical analysis (Euclidean distance, Ward’s method). AML-MP2 (a ∼75-79-year-old female), clustering with MPAL, had therapy related history for breast cancer but presented with T-ALL in lymph nodes and T/myeloid leukemia in the marrow. Genetics showed normal karyotype and *JAK3/NRAS* mutations, and therefore best classified as MPAL rather than (AML-MP/t-AML). **E.** Selected RNA-seq gene expression stratified by disease type. Among the genes upregulated in AML-MP and AML without MP samples were several master regulators of HSC such as *MEIS1*, *HOXB2*, and *HOXB3*.

To determine the specific lineage features separating MPAL from AML-MP, pathway analysis was performed using custom hematopoietic lineage gene sets derived from a published set of flow sorted hematopoietic stages derived from normal bone marrow samples^26^. Genes which were characteristic of each stage were identified using the Kruskal Wallis test statistic. Enrichment analysis using these sets revealed a significant HSC gene set enrichment in the AML-MP samples compared to MPAL in both myeloid (p< 0.001) and T blasts (p< 0.001) (**Figure 5B**). In comparison, monocytic and CD4 T-cell lineages were enriched in myeloid blasts (p<0.001) and T-blasts (p<0.001) sorted from MPAL, respectively.

GSEA using published gene sets in MSigDB^27,28^ revealed significant enrichment in gene sets characterized by leukemic stem cells (**Figure 5C**). GSVA scores were calculated for these gene sets and compared: the results highlighted a separation between MPAL and AML samples both with and without MP (**Figure 5D**). Among stem cell expression gene-sets, AML-MP samples had higher GSVA scores compared to MPAL cases. When comparing GSVA scores, AML-MP samples aggregated with AML without MP samples rather than MPAL. Sample-specific pathway enrichments using selected AML and hematopoietic stem cell gene sets revealed a greater enrichment of hematopoietic stem cells,^29^ Valk cluster 15^30^, upregulation of targets of NUP89/HOXA9 fusion in CD34+ hematopoietic cells,^31^ and upregulated genes in leukemic stem cells^32^ in AML and AML-MP samples. Unsupervised analysis of these enrichment scores identified two main sample groups: one composed entirely of AML and AML-MP samples, and the other composed almost entirely of MPAL samples (**Figure 5D**). To identify the genes driving this separation, the genes composing these pathways were examined individually (**Figure S5**). Among the genes identified were several master regulators of hematopoiesis such as *MEIS1*, *HOXB2*, and *HOXB3* (**Figure 5E**). Notably, one patient of AML-MP clustered with MPAL, displaying that at transcriptomic level it was closer to MPAL than AML-MP (**Figure 5D-E**). Therefore, this patient was biologically closer to MPAL despite technically meeting criteria of t-AML.

### AML-MP has limited lymphoid lineage plasticity with lower expression levels of lymphoid transcription factors

At diagnosis, the proportion of myeloid blasts in AML-MP was higher than that in MPAL (61%±35% vs 39%±37%, p=0.009, Wilcox test), while the portion of lymphoid blasts was comparable (Figure 6A). A subset of patients exhibited lineage shift from diagnosis either post-treatment or at a relapse event (**Figure 6A-C**). 7 AML-MP and 13 MPAL patients with persistent disease exhibited lineage shifts after induction therapy. Of the 24 patients who obtained complete remission but later relapsed, 8 (5 AML-MP, 3 MPAL) switched immunophenotypes between diagnosis and relapse and 6 of them became myeloid only. Interestingly, 3 patients had second relapse and the immunophenotype returned from myeloid only back to the original mixed phenotype. Altogether, among the patients remaining positive for disease after treatment (either post induction or relapse), comparable portion of MPAL and AML-MP patients displayed a myeloid- only immunophenotype (4/28, 14.2% vs 9/40, 22.5%, p=0.5, **Figure 6D**). Unexpectedly, MPAL patients frequently shifted to lymphoid (either B- or T-lineage)-only blasts (10/28, 35.7%), while patients with AML-MP rarely did (1/40, 2.5%, p=0.0003), suggesting a biological difference in *bona fide* lymphoid differentiation potential between AML-MP and MPAL.

**Figure 6:**
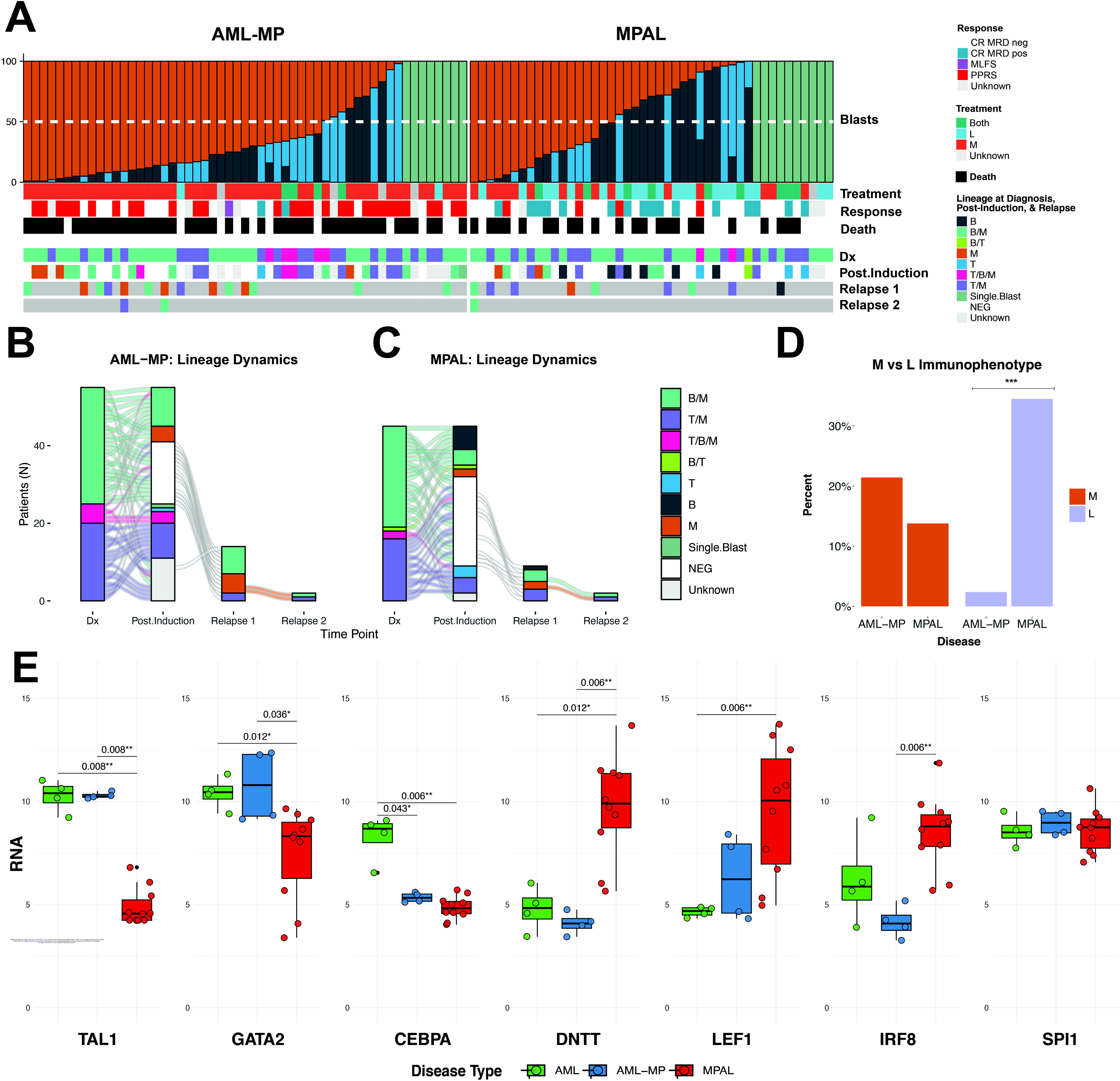
Limited lymphoid potential of AML-MP. **A.** Heatmap split by disease type, depicting blast percentages alongside blast immunophenotype per time point, treatment types and responses. Dx, diagnosis; CR MRD, complete remission, measurable residual disease; neg, negative; pos, positive; MLFS, morphologic leukemia free state; PPRS, partial, persistent, or refractory response. At relapse 1 or 2, grey boxes indicate patients either with death or no detectable disease. **B and C.** Alluvial plots per disease type exhibiting shifts in lineage between diagnosis, treatment, and relapse time points. **D.** Bar graphs comparing lineage shift post treatment, either post induction or relapse. M, myeloid blasts; L, lymphoid blasts. **E.** Floating bar charts depicting mRNA expression levels of select transcription factors critical for HSC, myeloid or lymphoid lineage commitment.

To provide mechanistic insights into diminished lymphoid differentiation potential in AML-MP, we evaluated mRNA expression levels of a list of transcription factors that are critical for myeloid versus lymphoid lineage commitment (**Figure S6**). Consistent with an enrichment for stemness signatures as shown by pathway analysis (see Figure 5), expression of *MEIS1*, *TAL1* and *GATA2*, all critical for HSC functions ^33–36^, was significantly higher in AML and AML-MP than in MPAL (**Figure 6E**). *SPI1* expression (encoding PU.1), a transcription factor indispensable for both myeloid and lymphoid commitment^37,38^, was comparable among 3 groups. I*RF8* mRNA, a master regulator for monocytic and dendritic lineage differentiation^39,40^, was higher in MPAL than AML-MP, consistent with the upregulated monocytic signatures (see Figure 5B). Notably, *CEBPA* mRNA, critical for myeloid differentiation^41^, was higher in AML while both low in AML-MP and MPAL. In contrast, mRNA expression of *DNTT* and *LEF1*, both involved in lymphoid commitment^42,43^, were significantly higher in MPAL than that in AML or AML-MP. In summary, AML-MP has higher levels of HSC transcription factors, while MPAL expresses higher levels of lymphoid transcription factors.

## DISCUSSION

Our study for the first time provides evidence that AML-MP clinically and biologically resembles its AML counterpart without MP but differs from MPAL, supporting the recommendation to exclude these patients from MPAL. This has important clinical implications as MPAL and AML-MP may be treated differently. Although there is no consensus on MPAL treatment, several retrospective studies have shown that induction therapy with ALL-type regimens is associated with superior outcomes particularly in pediatric patients,^6–10^ while a recent study from our group demonstrated 94% (16/17) patients with MPAL achieved complete remission after receiving a hybrid myeloid and lymphoid induction regimen.^44^ In contrast, the standard induction regimen for patients with AML-MRC/t-AML is liposomal formula of cytarabine and daunorubicin and more recently azacytidine and venetoclax.^11,45^ Our data suggests that patients with AML- MP may be best managed as AML-MRC/t-AML rather than MPAL, although novel therapies are urgently needed to improve the outcomes of both entities.

Our study has demonstrated the importance of excluding other entities from the diagnosis of MPAL and has also better defined the boundary between AML-MP and MPAL. First, patients with *RUNX1* (with or without other MR) mutations but otherwise manifesting as MPAL, behave similarly to the remaining MPAL patients, indicating that the presence of *RUNX1* mutations, contradictory to the recommendation by ICC2022, may not justify these MPAL to be reclassified as AML-MP. More data are needed to elucidate the implications of other MR mutations in this regard.^46^ Second, patients with driver fusions should be classified as such even if there is co-existing complex karyotype and/or *TP53* mutations. Our well annotated cohort study showed such adverse features (e.g. complex karyotype) typically seen in AML may not confer inferior outcome in *bona fide* MPAL, in contrast to a recent report.^47^ Third, ICC2022 classification removed therapy-related AML as an entity, therefore, how to classify therapy-related AML with mixed phenotype warrants further scrutinizing. Such patients with TP53 mutations and monosomal/complex karyotype may be best classified as AML with TP53 mutations. This notion is supported by our data showing lack of TP53 mutations as driver in MPAL. Conversely, in patients considered as therapy-related AML based on treatment history but with neither TP53 mutations nor MDS-CG, the classification of mixed phenotype requires comprehensive evaluation on a case-by-case basis. The data from our study indicate that the presence of nodal disease and/or mutations enriched for T-lymphoblastic lymphoma/leukemia (e.g. *NOTCH*, *JAK3*) may favor a diagnosis of MPAL in spite of prior cytotoxic therapy.

A unique finding of this study is the clusters identified by our novel unsupervised hierarchical clustering approach based on raw immunophenotypic files. Despite of the limited protein markers available (particularly no T-cell markers), this unbiased automated computational analysis accurately revealed immunophenotype, genotype and disease type correlation in certain clusters including cluster 5 as T/M phenotype, validating the utility of this approach. A recent study applied a similar approach to transcriptomes of a large cohort of so-called MPAL patients and identified 8 distinct clusters with immunophenotype and genotype correlation.^16^ G1-G4 with T/M phenotype and G5-G8 with B/M phenotype. Interestingly, G2 and G3 were enriched for *NOTCH1* mutations but G2 had *DNMT3A* and G3 had *PHF6* mutations, two molecular subsets initially reported by our group.^14^ G4 had FLT3 mutations and high expression level of *BCL11B*, consistent with a recently identified *BCL11B* rearranged T/M subset.^48,49^ Intriguingly, G1 was exclusively *CEBPA* mutated with frequent *WT1* and *FLT3/RAS* mutations, likely representing bZIP *CEBPA* mutated AML, which often upregulates T-cell makers and can be misclassified as T/M MPAL.^50,51^ Of note, *CEBPA* mutation is rare (2.7%) in our MPAL cohort. G8 harbored *BCR::ABL1* fusions and high frequency of *RUNX1* co-mutations, a feature well described in CML BP, but rarely seen in MPAL.^52,53^ As the inclusion criteria of our cohort are different with exclusion of CML BP (G8) and bZIP *CEBPA* mutations (G1) while including AML-MP and AML, a comparison between the two studies is difficult, if not impossible. However, G2 likely corresponds to cluster 5 in our study as both are T/M enriched for *DNMT3A* mutations. Despite of these caveats, both studies demonstrate that unsupervised hierarchical clustering based on either transcriptome or surface protein expression is a powerful tool to study these diseases with a high degree immunophenotypic and genetic heterogeneity. Our data also show that although the majority of AML-MP is clustered differently from MPAL, there is an overlap between these two cohorts of patients, which may be due to limited protein markers in our flow panels and/or represent a real biological overlap. In future, an integrated approach combining FCS files, transcriptomes, chromatic accessibility, and genetics of a large cohort of AML-MP and MPAL may be required to further delineate these patients.

The observation that AML-MP has diminished lymphoid differentiation potential is intriguing. This implies that AML-MP and MPAL differ in biology despite share immunophenotypic heterogeneity. At mutational levels, MPAL lacks TP53 mutations and many (although not all) MR gene mutations. Our gene set pathway analysis showed upregulation of several HSC, leukemic stem cell and AML gene sets in AML-MP and AML without MP. The expression of genes that regulate HSC and leukemic stem cell biology such as *TAL1*, *GATA2*, *MEIS1*, *HOXB2*, and *HOXB3.*^33–36,54,55^ was also upregulated in AML-MP and AML without MP as compared to MPAL. Thus, primitive/HSC differentiation program is a shared feature between AML-MP and AML without MP, further supporting classifying AML-MP as AML rather than MPAL. In contrast, MPAL showed increased expression of transcription factors critical for lymphoid development. Consistent with our findings, previous studies using lineage deconvolution have shown that primitive/HSC signatures are enriched in *TP53* and *RUNX1* mutated AML and are associated with inferior outcomes.^56,57^ In our small cohort of transcriptomes, a few patients with AML-MP grouped with MPAL, raising the possibility that the differential diagnosis between these two entities solely based on clinical history may not be perfect. Indeed, one of the AML-MP patients did not show HSC/MPP signatures or HSC gene set enrichment. Review of this patient’s clinical and genetic data supported a diagnosis of MPAL rather than AML-MP, illustrating the additive value of transcriptome to the classification.

The genetic and epigenetic mechanisms for mixed phenotype in AML-MP remain unclear. *RUNX1* mutations were present in 40% of AML-MP, two times more common than AML without MP, implicating a role of *RUNX1* mutations in this process.^58^ *RUNX1* has been shown to play an important role in lineage specification in hematopoietic and nonhematopoietic system.^59–65^ *RUNX1* mutations lead to megakaryocytic differentiation block but increased plasmacytoid dendritic cell differentiation.^65,66^ As *RUNX1* largely functions as a suppressor, loss-of-function mutations may result in imbalance of lineage specific (i.e. lymphoid) gene expression. The exact mechanism of how mutated *RUNX1* regulates lineage commitment awaits future studies. On the other hand, several studies have shown that immunophenotypically distinct blast compartments of MPAL harbor nearly identical genetic profile within a patient, and that cells from one lineage can reconstitute the alternate lineage in xenograft models,^13,14^ suggesting that other factors beyond genetics may also contribute to phenotypic heterogeneity. Future studies combining lineage tracing and single cell multi-omic technology will provide better insights into this intriguing biological process.^67,68^

## Data Availability

All data produced in the present study are available upon reasonable request to the authors

## Acknowledgement

This study was funded in part through the NIH/NCI Cancer Center Support Grant P30 CA008748. P.G. and A.C. are supported by grants from Department of Pathology and Laboratory Medicine at MSKCC. R.L.L. is supported by a Cycle for Survival Innovation Grant and National Cancer Institute R35 CA197594. J.L.G. is supported by a National Cancer Institute grant (NCI K08CA230172) and Equinox Cycle for Survival. WX is supported by Alex’s Lemonade Stand Foundation and the Runx1 Research Program, a Cycle for Survival’s Equinox Innovation Award in Rare Cancers, MSK Leukemia SPORE (Career Enhancement Program, NIH/NCI P50 CA254838) and a National Cancer Institute grant (K08CA267058).

## Authorship contributions

P.G., A.C., M.R. and W.X. identified patients and reviewed pathology. D.D. and J.G. performed automated flow cytometric analysis, analyzed RNA-seq data, and plotted figures. Y.Z. reviewed cytogenetic data. A.DerKach. performed statistical analysis. S.P., T.M. and Q.G. helped sorting. Y.L. and M.A. helped mutational analysis. A.Dogan. and R.L.L. supervised the project. W.X. and J.G. initiated, designed, and supervised the study. P.G., D.D., J.G. and W.X. wrote and all authors approved the manuscript.

