## supplementary Figures for "Acute myeloid leukemia with mixed phenotype is characterized by *RUNX1* mutations, stemness features and limited lineage plasticity"

Running title: Acute myeloid leukemia with mixed phenotype

Pallavi Galera^1,*^, Deepika Dilip^2,*^, Andriy Derkach^3^, Alex Chan^1^, Yanming Zhang^4^, Sonali Persuad^5^, Tanmay Mishera^5^, Ying Liu^1,6^, Christopher Famulare^7^, Qi Gao^1^, Douglas A. Mata^6^, Maria Arcila^1,6^, Mark B. Geyer^8^, Eytan Stein^8^, Ahmet Dogan^1^, Ross Levine^2,5,7,8^, Mikhail Roshal^1^, Jacob Glass^2,7,8^, Wenbin Xiao^1,7^

**Methods**

**Chromosome and FISH analysis**

Conventional chromosome analysis of bone marrow, peripheral blood, or fresh tissue was performed following standard procedures with overnight short-term culturing without mitogen. At least 20 metaphase cells were analyzed for a complete chromosome study. The chromosome abnormalities were recorded as per the International System for Human Cytogenetic Nomenclature (2016). In most cases, fluorescence in situ hybridization (FISH) analysis was performed for recurring chromosome abnormalities in AML and ALL, including t(9;22), MLL(11q23), and t(12;21). In a few cases with inadequate chromosome analysis, extensive FISH tests for deletion 5q31, deletion or loss of chromosome 5, trisomy 8, deletion 7q31, deletion or loss of chromosome 7, deletion 17p13.1 (TP53), deletion or loss of chromosome 17, 3q36 rearrangement, deletion 16q22, deletion 20q12, t(8;21), inv(16), t(6;9), and IKZF1 were also performed. All FISH probes were from Abbott Molecular (Des Plaines, IL), and quality and performance were validated in the laboratory. All the cytogenetic results were re-annotated by a board-certified cytogeneticist (Y.Z.)

**Flow cytometry and cell sorting**

Multiparameter flow cytometry was performed on bone marrow aspirates at diagnosis. Briefly, up to 1.5 million cells from freshly drawn bone marrow aspirate were stained with 10-“color” panels (**Supplemental Table S1**)^28^, washed, and acquired on a Canto-10 cytometer (BD Biosciences, San Jose, CA). The results were analyzed with custom Woodlist software (generous gift of B. L. Wood, University of Washington). Cryopreserved bone marrow aspirate specimens collected at diagnosis from our institutional, IRB- approved biospecimen bank were thawed and flow sorted into myeloid blasts and T lymphoblasts based on the immunophenotype using FACS-Aria Fusion cell sorter (BD Biosciences)^14^.

**Sequencing studies**

Bone marrow samples obtained were submitted to a 28-gene capture-based next-generation sequencing (NGS) assay (RainDance) or a larger panel with over 400-gene capture-based NGS assay (FoundationOne Heme or MSK-IMPACT) as previously described^29-31^. In addition, an RNA-based targeted sequencing assay (Archer Fusion- Plex Heme Kit) that detects and identifies fusions of 87 genes associated with hematological malignancies was performed. Variants were detected through our clinical workflow, which includes sequencing by Raindance or IMPACT custom panels and FoundationOne Medicine..

**Hierarchical clustering based on flow cytometry data**

Raw flow cytometry data was obtained in FCS file format. A flow gating pipeline was used to process flow cytometry standard (FCS) files. Following compensation, the samples were subsetted using gates for singlets, viable cells, and dim CD45 expression for both the ‘M1’ and ‘M2’ myeloid sample tubes (**Supplemental Table S1**). A 20 x 20 self-organizing map (SOM) was then used to identify cell clusters in each tube in the merged dataset, resulting in a combined total of 400 clusters. Cell counts in each cluster were then tallied for each sample and adjusted using a variance-stabilizing transformation. Hierarchical clustering using Ward’s method and Euclidean distance was used for unsupervised analysis of the samples as well as a principal component analysis. Clinical annotations, mutation data, and pathologist assigned immunophenotypes were also integrated. The code developed for this automated flow file analysis is available upon request.

A principal component analysis (PCA) was used to assess the variance between the samples based on FCS data. A PCA was performed on M1, M2, and M1 and M2 combined data. Samples were designated per disease type, oncoprint FCS cluster, and immunophenotype. Immunophenotypes were tabulated per FCS cluster to determine if specific FCS clusters reflected immunophenotypes.

**Pathway Analysis of transcriptome**

Gene sets corresponding to the stages of hematopoietic differentiation were constructed from genes in the Corces peak atlas^32^. Genes differentially expressed between the MPAL, and AML-MP cohorts were identified for both M and T subsets in parallel using DESeq2^33^. Genes that contained a p-adjusted value of 0.05 or less and a log fold change of 1.5 or higher were used for quantifying enrichment. Enrichment for specific hematopoietic stages was assessed using gene-ontology over-representation analysis with up and downregulated genes analyzed separately^34^.

Gene set enrichment analysis (GSEA) was also performed to identify pathways distinguishing MPAL from AML-MP cases. Sample specific enrichments for these sets were calculated using GSVA. Hierarchical clustering of these sample specific pathway enrichments was performed using Euclidean distance and Ward’s method. Hierarchical clustering was also performed on the expression data for the individual genes in these pathways. Additionally, genes from the Valk geneset^35^ and an LSC geneset^36^ were used to visualize contrasts in RNA-seq expression levels. Row clustering was applied, and genes that drove contrasts between disease groups were visualized. Pairwise contrasts through DESeq2 were also ran between disease groups to determine statistical significance.

**Statistical analysis**

Patient characteristics were summarized by frequency (percentage) for categorical. Associations between MPAL, AML-MP, AML without MP and patient and were tested by Fisher’s exact test. Associations with continuous characteristics were evaluated by Wilcoxon rank-sum test. The overall survival (OS) was evaluated by Kaplan-Meier method and the difference between groups was determined by log-rank test. The effects of patient and disease characteristics on OS were estimated by univariate Cox proportional hazard model with p<0.05 being considered significant. All statistical analyses were performed using R.

**Supplemental Figure 1.**

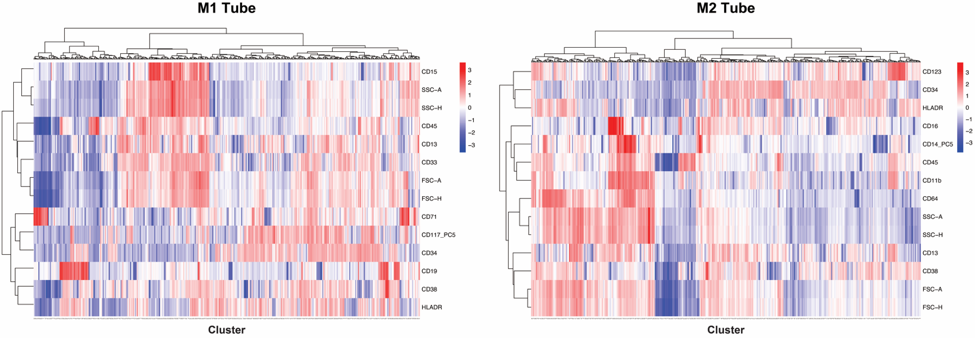

**Figure S1. Self-organizing map (SOM) to identify immunophenotypically distinct populations.** Flow cytometric data following compensation was subsetted using gates for singlets, viable cells, and dim CD45 expression for M1 and M2 myeloid tubes (supplemental Table 1). A 20 x 20 SOM was then used to identify cell clusters in each tube in the merged dataset that identified 200 distinct populations in the combined cohort. The clusters were subsequently combined to create a composite 400 feature classifier for the samples in the cohort. Hierarchical clustering is performed here for visualization purposes using Ward’s method and Euclidean distance.

**Supplemental figure 2.**

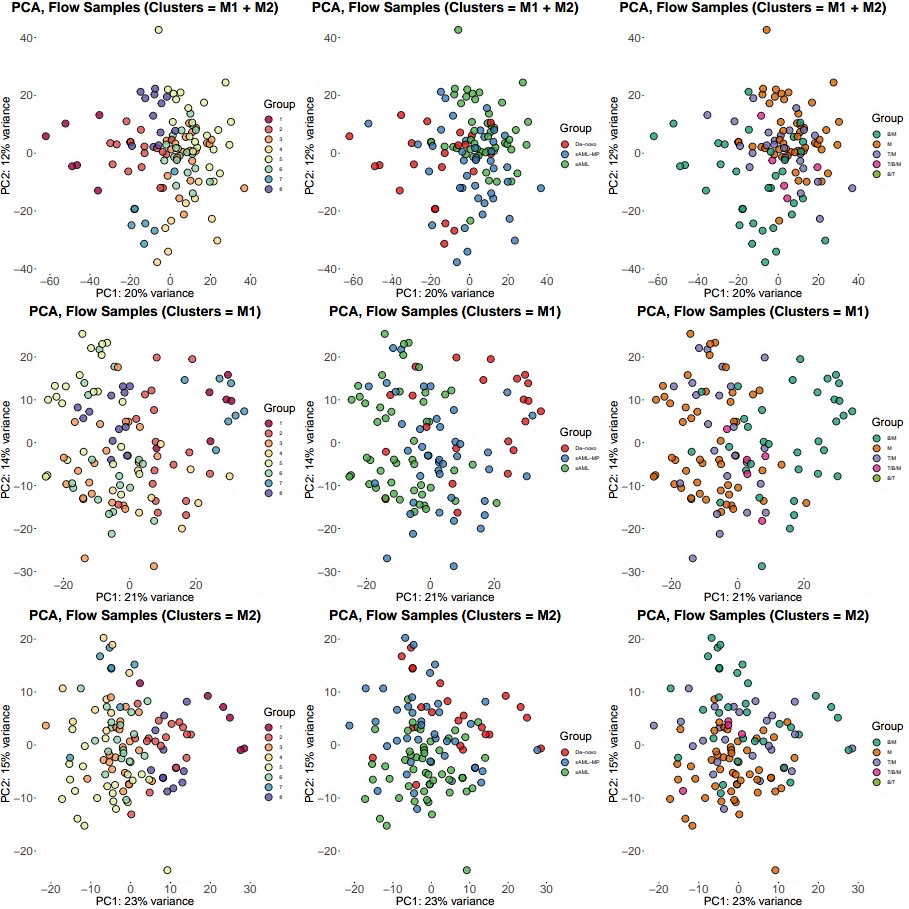

**Figure S2. Principal components analysis (PCA) conducted on VST-transformed flow cytometry clusters.** PCAs were performed on flow cytometry clusters using M1, M2, and combined M1 and M2 tubes. Cases were colored by cluster identity, disease type, and lineage (B/M, M, T/M, T/B/M, B/T).

**Supplementary Figure 3**:

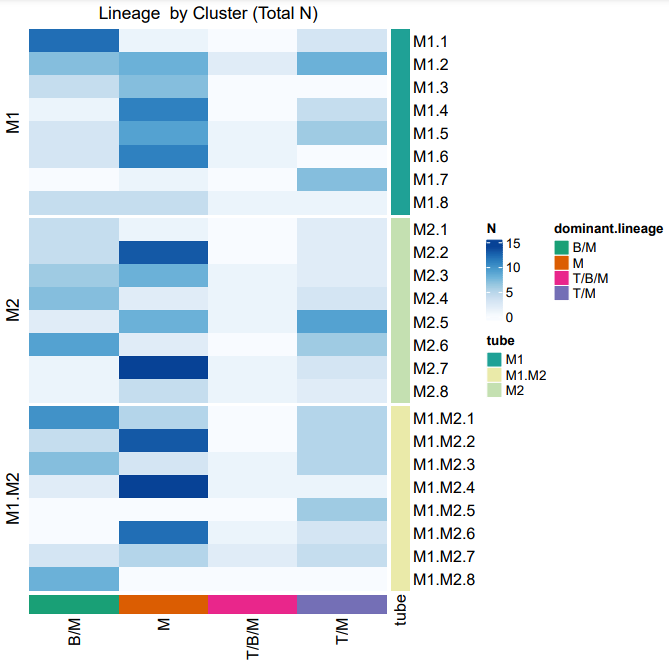

**Figure S3. Proportion of cases** **by immunophenotype in M1, M2, and combined M1/M2 tubes.** Rows are split by tube, with immunophenotype highlighted in the bottom most row (B/M, M, T/B/M and T/M).

**Supplementary Figure 4:**

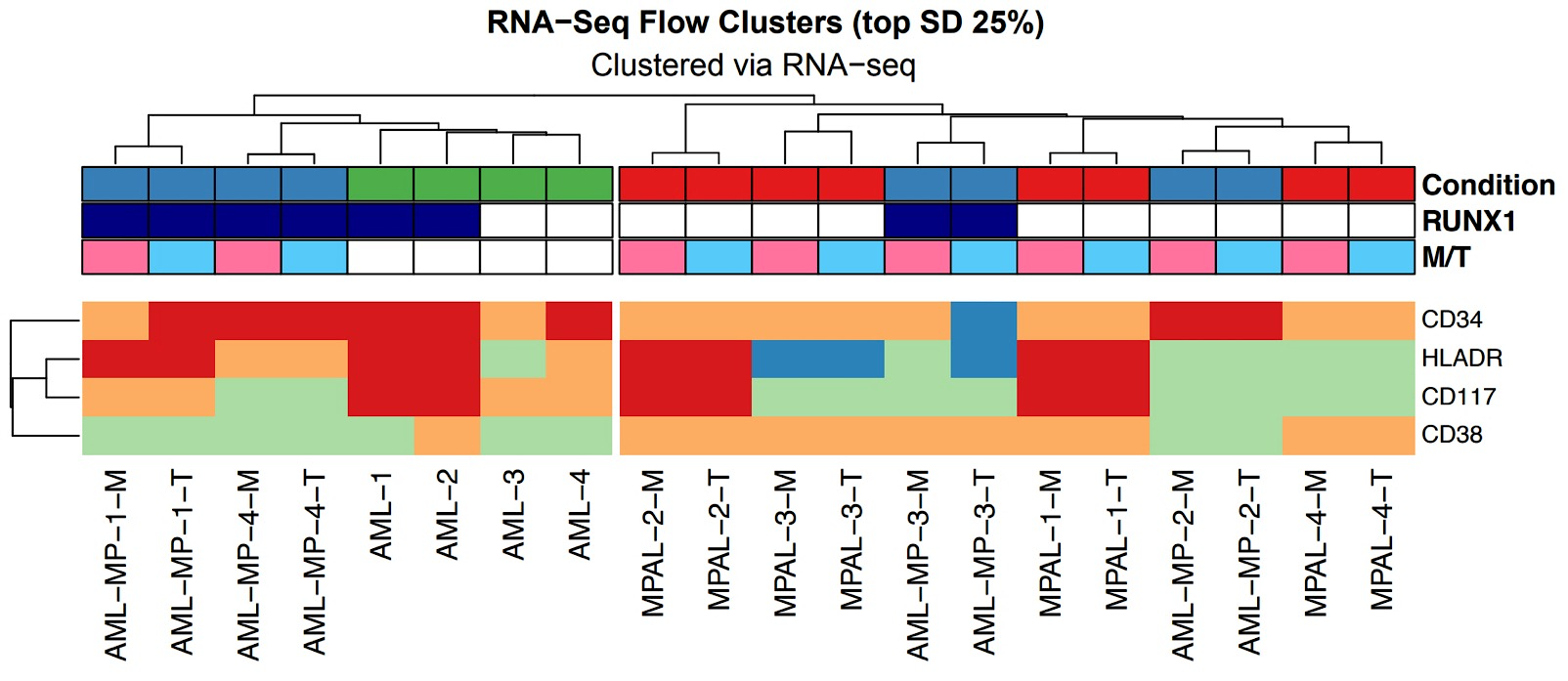

**Figure S4. The correlation between RNA-sequencing gene expression and flow cytometric surface maker expression.** Hierarchical clustering (Ward’s method, Euclidean distance) on VST-transformed RNA-sequencing data, annotated flow cytometry data and disease type. Cases are split into 2 distinct groups, with clustering highlighting differential hematopoietic catheterization and RUNX1 mutations.

**Supplementary Figure 5:**

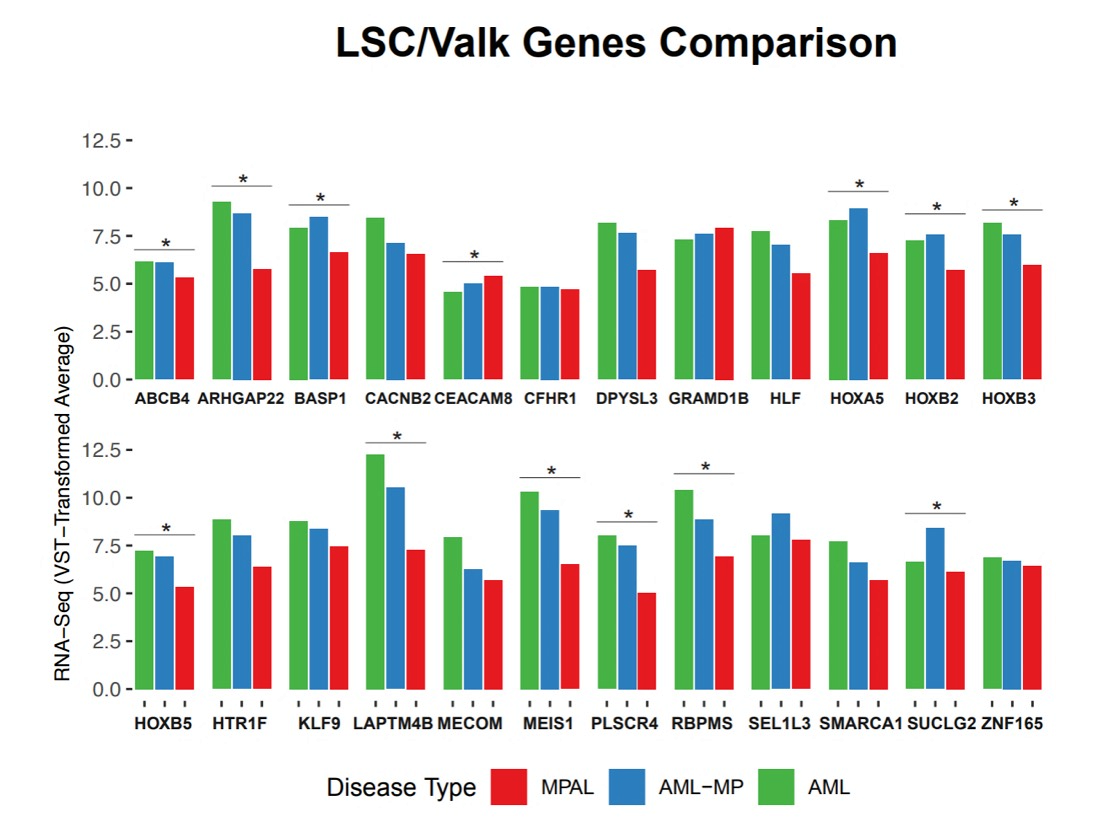

**Figure S5. Gene expression in AML-MP.** Bar graphs comparing RNA-seq VST-transformed counts per selected genes, annotated with statistical significance bars (p < 0.05).
